# DETERMINANTS OF SECOND CHILD BIRTH INTERVAL AMONG WOMEN OF LUSAKA PROVINCE: A COX REGRESSION MODEL

**DOI:** 10.1101/2023.02.15.23285966

**Authors:** M Zambwe, M Zambwe, R Mutemwa

**Affiliations:** School of Medicine and Health Sciences, University of Lusaka, Lusaka, Zambia; Benefits Department, Workers Compensation Fund Control Board, Lusaka, Zambia

## Abstract

**Background:** Child birth intervalis the length of time between two successful live births.Shorter child birth interval among women of reproductive age is a serious global public health challenge. It is associated with low birth weight, child malnutrition, and child mortality.There is a general dearth of literature on this subject in Zambia. Therefore, the aim of this study was to establish the determinants of second child birth interval among women.

**Methods:** A cross-sectional study involving 100 women of reproductiveage group in Lusaka province. The participants were purposively and convenientlyselected. A pre tested structured questionnaire was used to collect the data. The key variables in the data were age, marital status, educational level, tribe, income. The data from the questionnaires were summarized into a Microsoft excel sheet, and cleaned for errors and duplications.Thereafter the excel sheet was exported to Stata version 14.2 for analysis.Kaplan-Meier and Cox regression analysis wereapplied.

**Results:** The p-value for age was 0.00 which was statistically significant. Within the age group 18-25 years the average second birth interval was 23 months. In the age group 26-44 years, the average second birth interval was 42 months. The reported p-value for income was 0.049 which was significant. The results also showed that those with income between K0-K350 had an average second birth interval of child of 27 months. Those with income of K3500-K16000, had an average second birth interval 69 months. Women in urban areas had a longer second birth interval than those in rural areas.

**Conclusion:** Age, income and location were found to be significantly associated with the incidence of a second birth. A lot of awareness activities on birth control are therefore recommended among young, low income and rural women.

## Introduction

Fertility is one of the key determinants of population growth. It is very important analyze fertility in order to plan for population control and evaluate available family planning programs.^1^ Fertility depends on a lot factors such as couple’s decisions, health, education and child birth interval.^2^Child birth intervalis the length of time between two successful live births.Shorter child birth interval among women of reproductive age is a serious global public health challenge. It has been found to be associated with high incidences low birth weight, child malnutrition, and child mortality.^3^Child birth interval of less than 17 months or more than 5 years is highly associated with maternal and child health problems.^4^

A study in Latin America revealed that shorter child birth interval to have a second child a second child increased parinatal outcomes. Another study in India revealed that short birth interval increased child mortality by 67%. A study in Bangladesh revealed that an increase in the birth interval improvised the child survival.

Demographic and Health Surveys (DHS) data from 18 developing countries in Asia, Latin America, Africa and the Middle East showed that a birth interval of three-years improve the survival status of under five children10. Another similar survey from 52 developing countries revealed that too short birth intervals are associated with adverse pregnancy outcomes, increased morbidity in pregnancy, and increased infant and child mortality.

Child birth interval beyond two years improves maternal health by decreasing maternal morbidities (such as toxaemia, anaemia and third trimester bleeding) and mortality. In sub-Saharan Africa, about 60% of women deliver the next child before the index child celebrates his/her third birth day and almost a quarter before the second birth day12. The child mortality rate in Zambia as of 2021 was at 61.4 deaths per 1000 live birth. Although there are enough studies on fertility and child spacing in Zambia, there is very little available prognostic models that establish the determinants of child birth interval. The aim of this study was therefore to establish the determinants of second child birth interval among women of reproduction age in Chipata district.

## Materials and methods

### Study Design

A cross□sectional study review of women in the age group 15 to 49 years domiciled in Lusaka Province was conducted.

### Study Population

The study population consisted of all women of reproductive age group in Lusaka province. Such women were either in schools, colleges, work places or at home.

### Inclusion Criteria

All women in the age group 15 to 49 years. Further those women need to have had a successful first child birth.

### Exclusion Criteria

The study excluded all females less than 15 years of age and those above 49 years. Further all females without record of a successful first child birth were excluded.

### Sampling Procedure

As the study was not funded a sample size of 100 participants was selected using purposive and convenience methods. Fifty participants were drawn from two rural districts of Lusaka province – Chongwe and Rufunsa. And another fifty was drawn from Lusaka urban district of the province.

### Ethical Approval

This work was approved by the University of Lusaka Ethics Committee, School of Medicine & Health Sciences Research Ethics Committee under reference: IORG0010092. Participation was voluntary. The aim of the study was explained to the participants prior to the interview after which written consents from them were obtained.

### Data collection

There were no interventions or treatments in this study. Primary data were drawn from the participants using a structured questionnaire. The validity of the questionnaire was confirmed by 10 senior demographers and sociologists.The data variables included age, education, income, marital status, location and tribe.

### Data Analysis

Data from the questionnaires were individually entered into a Microsoft excel sheet. The data were checked for duplications and errors and then finally exported to Stata Version 14.0 for analysis. Categorical variables were summarized using proportions, while continuous variables were summarized using means and standard deviations as appropriate. The key dependent variable was the second child interval. The effect of independent variables on the dependent variable was investigated using cox regression and the Kaplan-Mier curve. The level of significance was set at 5% (p < 0.05).

The cox proportional hazard model was applied because it investigates the effects of the covariates on the second birth interval which was the aim of the study (Cox, 1972).^5^

The mathematical form of hazard function for the Cox proportional hazard model is:

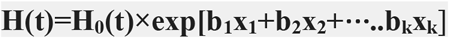

Where *x*_1_ ⋯ *x*_*k*_ *are* factors that influence the time to event: *H*_0_(*t*) is the baseline hazard at time *t*, this is the Hazard of an individual having predictors at time zero. By computing the exponential of the regression coefficient “*b*_1_ ⋯ *b*_*k*_” (directly provided by the software), the HR of a given predictor in the model is determined.

This expression gives the hazard rate at time t for the subject i with covariate vector (explanatory variables) Xi. The usual Cox proportional hazard model requires time-to-event data to be independent (Sighn, 2011).^6^

The event of interest was the time to second-birth interval in months.

## Results

**Table 1:**
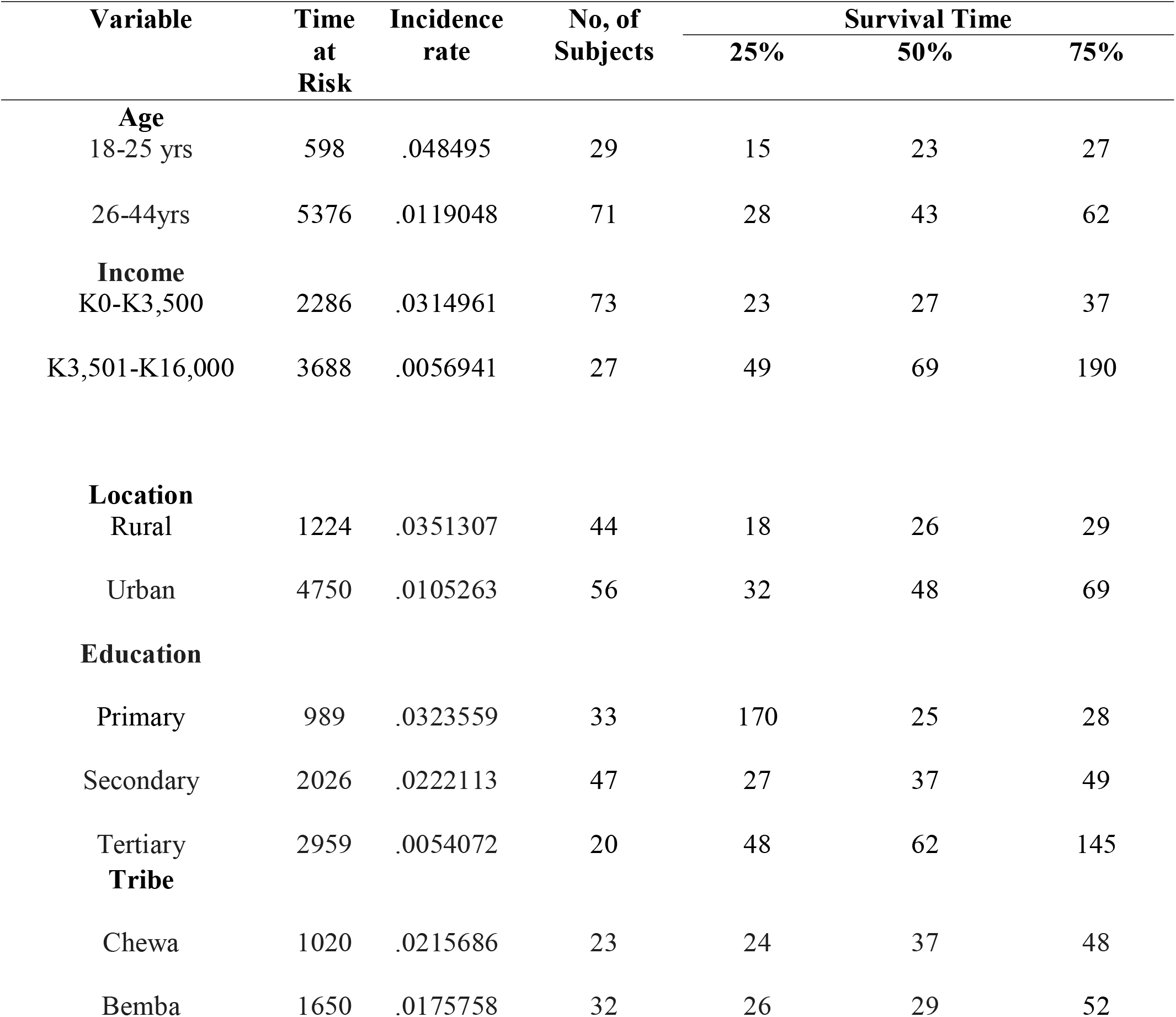
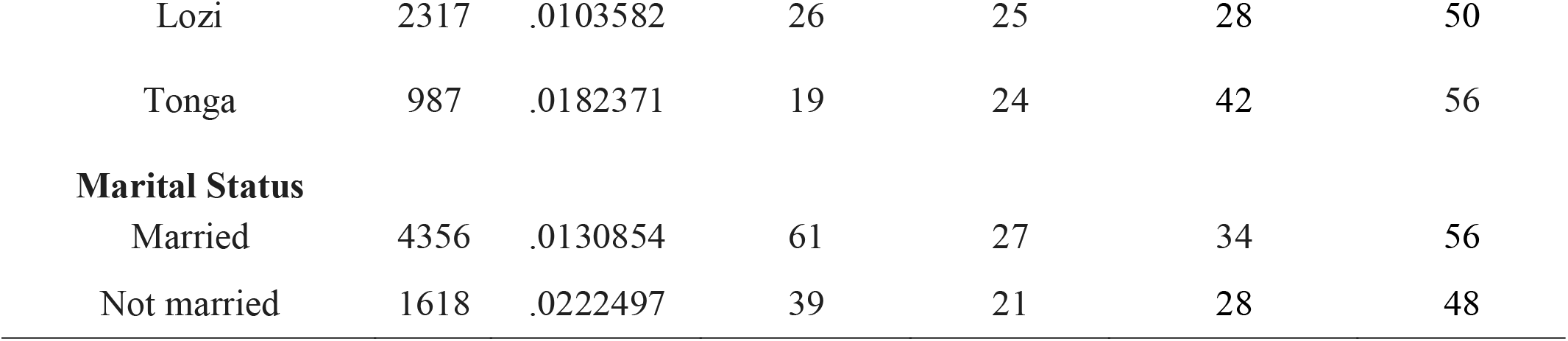
Second Child Interval Socio-demographic characteristics of women participants in Lusaka province, Zambia 2022. (n=100)

| Variable | Time<br>at<br>Risk | Incidence<br>rate | No, of<br>Subjects | Survival Time |  |  |
| --- | --- | --- | --- | --- | --- | --- |
|  |  |  |  | 25% | 50% | 75% |
| **Age** |  |  |  |  |  |  |
| 18-25 yrs | 598 | .048495 | 29 | 15 | 23 | 27 |
| 26-44yrs | 5376 | .0119048 | 71 | 28 | 43 | 62 |
| **Income** |  |  |  |  |  |  |
| K0-K3,500 | 2286 | .0314961 | 73 | 23 | 27 | 37 |
| K3,501-K16,000 | 3688 | .0056941 | 27 | 49 | 69 | 190 |
| **Location** |  |  |  |  |  |  |
| Rural | 1224 | .0351307 | 44 | 18 | 26 | 29 |
| Urban | 4750 | .0105263 | 56 | 32 | 48 | 69 |
| **Education** |  |  |  |  |  |  |
| Primary | 989 | .0323559 | 33 | 170 | 25 | 28 |
| Secondary | 2026 | .0222113 | 47 | 27 | 37 | 49 |
| Tertiary | 2959 | .0054072 | 20 | 48 | 62 | 145 |
| **Tribe** |  |  |  |  |  |  |
| Chewa | 1020 | .0215686 | 23 | 24 | 37 | 48 |
| Bemba | 1650 | .0175758 | 32 | 26 | 29 | 52 |
| Lozi | 2317 | .0103582 | 26 | 25 | 28 | 50 |
| Tonga | 987 | .0182371 | 19 | 24 | 42 | 56 |
| **Marital Status** |  |  |  |  |  |  |
| Married | 4356 | .0130854 | 61 | 27 | 34 | 56 |
| Not married | 1618 | .0222497 | 39 | 21 | 28 | 48 |

The Kaplan-Meier curve on figure1 shows that the women in rural areas (area 1) have a shorter time taken to have a second child compared to those in the urban area (area 2).

**Fig 1:**
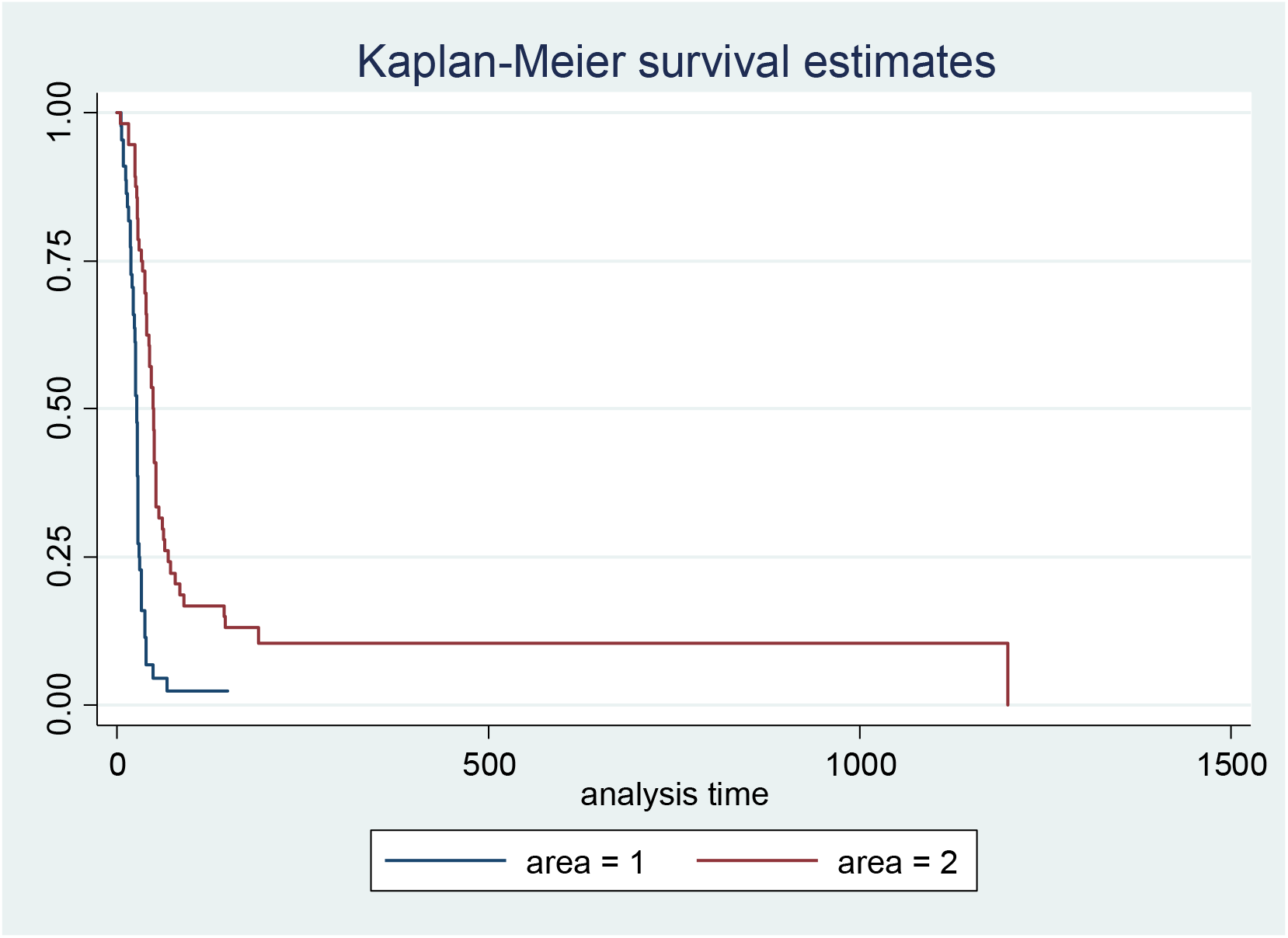
Kaplan-Meier curve depicting second birth interval of Lusaka women in rural and urban areas.

**Table 2.**
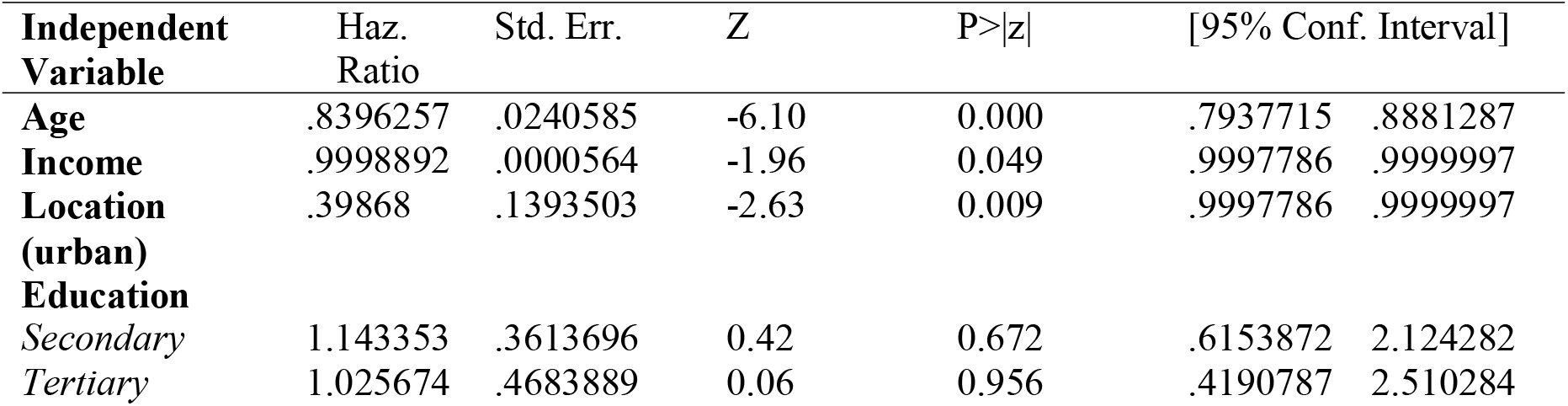
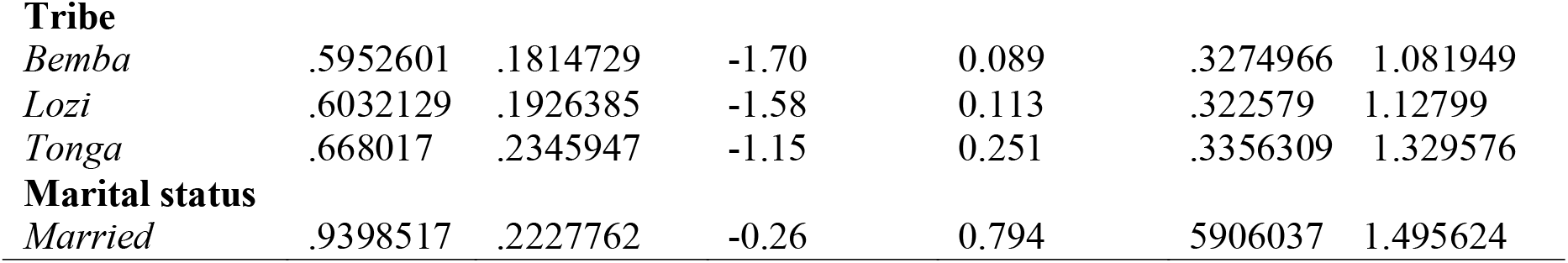
Cox regression analysis on time to second-child in women of Lusaka province.

### Age

For every unit increase in age, the hazard of having a second child birth in women was 0.84 or 16% less. This is statistically significant because the P-value was found to be 0.00 which is below the 0.05 significant levels.

### Income

For every unit increase in income the hazard of having a second child birth was 0.998 or 11% less. This was statistically significant because the P-value is 0.049 which is below the 5% significant level.

### Location

The hazard of having a second child birth for women in the urban areas was 0.3986 or 60% lower compared to those in the rural area. The P-value was found to be 0.009 which is statistically significant as it is below the 5% significant level. At 95% confident interval we can as well say that the hazard of having a second child birth for the women in a true population in urban area was 0.2 lower limit and 0.79 upper limit.

### Education

The hazard of having a second child birth in the women with a secondary level education was 1.14 or 14% greater when compared to those that had a primary level education. However, this was not statistically significant because the P-value was greater than the 5% significant levels. Therefore, there was no association between having a second child birth and primary or secondary education status. The hazard of having a second child birth in the women with a tertiary education level was 1.02256 times greater when compared to the people to the people with a primary education level. This was not significant because the p-value was greater than the 5% significant level.

### Marital status

The hazard of having a second child birth in women that are not married was 0.93 or 7% greater when compared to those that were married. This was not statistically significant because the p-value was found to be greater than 5%. There was therefore, no association in the hazard of having a second child in the married and those that were not married.

### Tribe

The hazard of having a second child birth for Bemba women was 0.5952 or 40% less when compared to a Chewa. This was not statistically significant because the p-value was above the 0.05 significant level. There was therefore no association or difference in the hazard of having a second child birthamong Chewa and Bemba women. The hazard of having a second child birth for Loziwomen was 0.60 or 40% less as compared to Chewas. This too is not significant because the p-value was 0.113 which is greater than the 5% significant level. The hazard of having a second child birth for a Tonga womanwas 0.668 or 33.2% less than the hazard of having a second child for Chewa. The p-value was 0.251, higher than the significance level, hence not significant.

## Discussion

To the best of our knowledge, there is a dearth of literature in Zambia, done to establish the timing between age of a women and the second birth. The aim of this study was therefore to establish the factors that determine the transition from first child birth to second child birth using a non parametrictest (Kaplam Meier curve) and semi-parametric test (Cox regression analysis). The results in this study revealed that the age, level of income and locationof a woman were significantly associated with the transition from first child birth to second child birth. The results showed that the older a women becomes the longer the timing to a second child. This finding is consistent with a similar study in Bangladesh.^3^The age of women at birth is very important as it determine the health status of the mother as well as the children respectively.

The results in this study also revealed that women with low income have a shorter transition from first child to second child as compared to those that had high income. The income level of income of women is very important in determining their reproductive health behavior. Women with higher income have access to contraceptives as compared to those with lower income. This findingis consistent with a similar study done in Bangladesh, where it was established that shorter birth internal in women is associated with low income.^7^

Our study revealed that women in urban areas have a longer transition from first birth to second birth as compared to the women in the rural areas. The living location of a woman is very important as it determines their access to health services like access to contraceptives. This finding was in agreement with another study in Ethiopia; where it was establishedthat that women in-rural areas have a shorter birth interval as compared to those that live in urban areas.^8^

On education, most studies in literature have established an association between it and child bearing. However, our study could not establish an association. We can just speculate that maybe our study was on second birth incidence. Whereas most studies focus on first birth incidence.

Our study had limitations. The participants in the study were not randomly selected, giving each member of the target population an equal chance to be part of the sample. Rather the participants were conveniently and purposively selected due to lack of financial resources. So the results can not be generalized to the entire population.

## Conclusion

Our study went out to establish the determinants of child birth interval among women of reproductive age in Lusaka province. Age, income and location were found to be significantly associated with the incidence of a second child.in view of the above, a lot of awareness and health promotion activities are recommended among young, low income and rural women in their fertility potentials to having more children.

## Data Availability

All data produced in the present study are available upon reasonable request to the authors

https://www.unilus.ac.zm/

